# Network Assisted Analysis of *De Novo* Variants Using Protein-Protein Interaction Information Identified 46 Candidate Genes for Congenital Heart Disease

**DOI:** 10.1101/2021.11.30.21267069

**Authors:** Yuhan Xie, Wei Jiang, Weilai Dong, Hongyu Li, Sheng Chih Jin, Martina Brueckner, Hongyu Zhao

## Abstract

*De novo* variants (DNVs) with deleterious effects have proved informative in identifying risk genes for early-onset diseases such as congenital heart disease (CHD). A number of statistical methods have been proposed for family-based studies or case/control studies to identify risk genes by screening genes with more DNVs than expected by chance in Whole Exome Sequencing (WES) studies. However, the statistical power is still limited for cohorts with thousands of subjects. Under the hypothesis that connected genes in protein-protein interaction (PPI) networks are more likely to share similar disease association status, we develop a Markov Random Field model that can leverage information from publicly available PPI databases to increase power in identifying risk genes. We identified 46 candidate genes with at least 1 DNV in the CHD study cohort, including 18 known human CHD genes and 35 highly expressed genes in mouse developing heart. Our results may shed new insight on the shared protein functionality among risk genes for CHD.

## Background

Congenital heart disease (CHD) is the most common birth defect affecting ∼ 1% of live births and accounts for one-third of all major congenital abnormalities [1-3]. There is substantial evidence that CHD has a strong genetic component [4]. Although it is estimated that aneuploidies and copy number variations account for about 23% of CHD cases, few individual disease-causing genes have been identified in published studies [5-8]. Therefore, the limited knowledge on the underlying genetic causes poses an obstacle to the reproductive counseling of CHD patients [9].

Whole Exome Sequencing (WES) studies have successfully boosted novel causal genes identification for both Mendelian and complex disorders [10, 11]. To narrow down the pool of candidate variants from WES, family-based studies have been conducted to scan for *de novo* variants (DNVs) from parent-offspring trios. DNV studies have been shown to play an important role in risk gene identification for CHD [1, 3, 5, 6, 12-15]. From the analysis of 1,213 CHD parent-offspring trios, Homsy et al. identified greater burden of damaging DNVs, especially in genes with likely functional roles in heart and brain development [12]. Recently, Jin et al. inferred that DNVs in ∼440 genes were likely contributors to CHD [5]. Despite these advances, it remains challenging to capture the causal genes with only DNV data as CHD is very genetically heterogeneous [6].

It is believed that genes interact with each other in biological processes and may jointly affect the disease risk through biological pathways. In recent studies on CHD, researchers reported significant enrichment of genes related to histone modification, chromatin modification, cilia function, transcriptional regulation, neural tube development, and cardiac development and enrichment in pathways including Wnt, Notch, Igf, HDAC, ErbB and NF-kb signaling [1, 3, 12, 14, 16]. These results suggest that considering functional relationships of genes through pathway-level analyses may complement gene-level analyses and improve risk gene identification for CHD.

Recently, Nguyen et al. proposed a framework to conduct pathway-level analysis on schizophrenia DNV data [17]. Their method is built on the extTADA framework and integrates pathway information by multiplying a gene-set related term in the likelihood function. However, this method requires curated pathways related to schizophrenia and treats genes as exchangeable without considering gene-gene interactions. To characterize the functional connectivity between genes, we can consider genes as a network by modeling the topology of pathways.

Network-based approaches have been successful in prioritizing risk genes for downstream analysis of Genome-Wide Association Studies (GWAS) and gene expression studies [18-20]. Chen et al. [19] proposed a Markov Random Field (MRF) model to incorporate pathway topology structure for GWAS. They showed that their method is more powerful than single gene-based methods through both simulation and real data analyses. Following the idea of this framework, Hou et al. [20] proposed a method that integrates co-expression networks and GWAS results. By applying their method to Crohn’s disease and Parkinson’s disease, they showed that their method could lead to more replicable results and find potential disease-associated pathways. More recently, Liu et al. adopted a similar idea as Chen et al. and Hou et al. to analyze DNV data from WES studies [21]. Their framework, namely DAWN, combines TADA p-values with estimated network from gene co-expression data. In their real data analysis for autism, 333 genes were prioritized by integrating DNV summary statistics and expression data from brain tissue. However, all of these above methods require summary statistics (Z scores or p-values) from genetic association analysis as their input, which may not be provided for DNV analysis results [22, 23]. In addition, it is difficult to apply DAWN directly to CHD because of the lack of ideal co-expression data from relevant tissues (such as developmental heart) for CHD.

As there is a limited number of co-expression data sets for human developmental heart, a natural choice for network information would be human protein-protein interaction (PPI) databases. There are multiple public sources of PPI network databases such as BioGRID [24], IntAct [25], DIP [26], MINT [27], and HPRD [28]. Most of network-based studies apply their real data on two or more of the databases to obtain their results. Nonetheless, it is hard to check the overlapping information between two PPI databases and interpret the divergent results. The STRING [29] database provides a platform to resolve the above problems. It imports protein association knowledge from physical interaction and curated knowledge from the aforementioned PPI databases and other pathway information knowledge such as KEGG and GO. In addition, it provides a score to measure the likelihood of interactions. Some studies have used STRING in their post-association analysis for gene-based DNV studies and showed significant enrichment of candidate CHD risk genes in the STRING PPI network [30, 31]. One recent study [3] constructed a priority score based on canonical pathways and PPI networks and identified 23 novel genes for CHD. These results suggest that incorporating PPI network information from STRING may identify additional risk genes with more biological interpretability.

In addition, in the post-association analysis of CHD DNV analysis from our previous multi-trait method M-DATA [32], we found that the number of edges formed by candidate CHD genes (47 edges, green line in Fig 1A) identified by M-DATA multi-trait model (33 genes with false discovery rate (FDR) q-values < 0.1) is far larger than the upper tail of the empirical distribution sampled from 33 randomly selected genes in the STRING V11.0 database (score threshold: 400) for 10,000 times (Fig 1A). This suggests that the candidate CHD genes are highly enriched in terms of their interactions in the STRING database. To further illustrate that PPI information may contribute to CHD gene discovery, we use the CHD result from M-DATA single-trait analysis and use the result of autism as a comparison. Fig 1B shows the number of edges formed by the top genes (ranked by FDR q-values) for CHD and autism from M-DATA single-trait model with a more stringent selection of PPI edges in the STRING database (score threshold: 950), respectively. To compare with the number of edges formed by randomly selected genes, we plot the 95% quantile of the empirical distribution sampled from random genes in the STRING v11.0 database (score threshold: 950) for 10,000 times as a baseline. When more than 25 top CHD genes are selected, the number of edges formed by these genes is significantly more than that from randomly selected genes, whereas the number of edges formed by the autism top genes did not differ much from randomly selected genes. This suggests top genes in CHD tend to be neighbors in the STRING PPI network.

**Fig 1.**
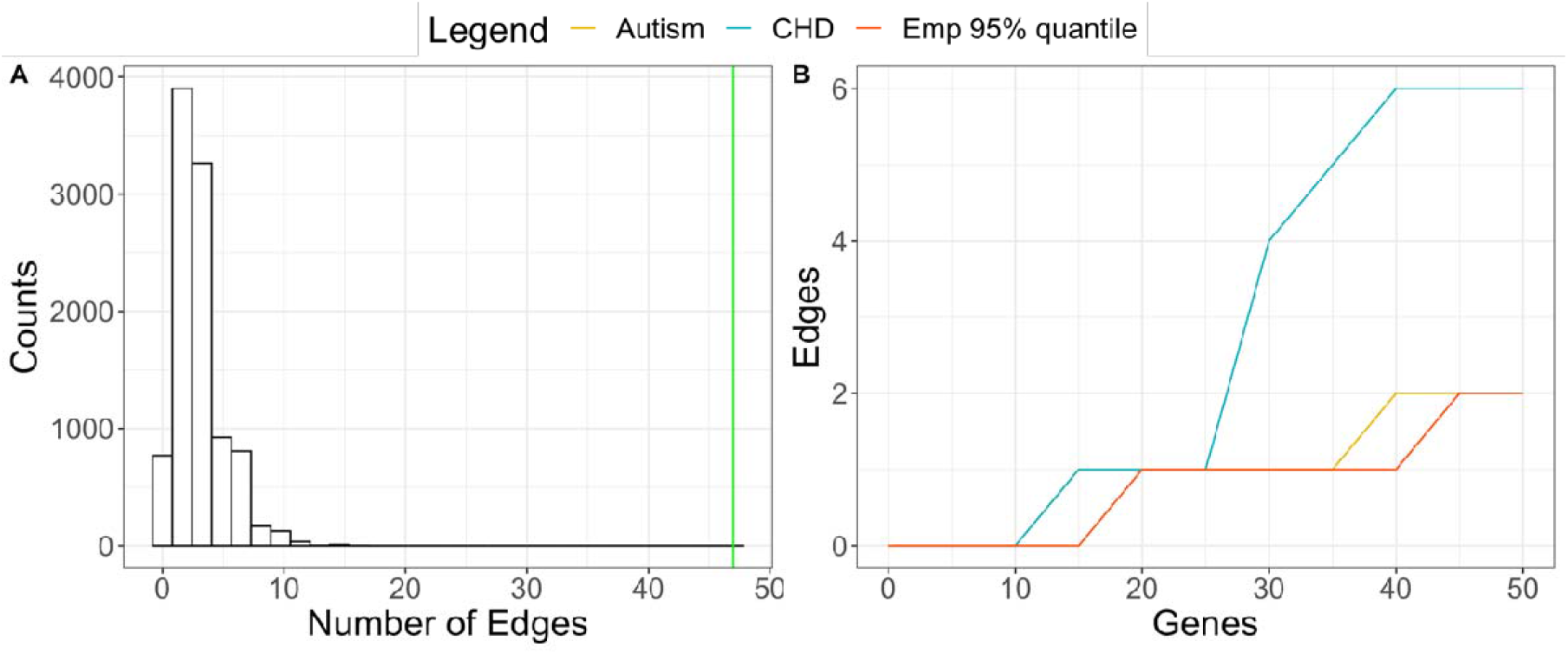
CHD top genes are more connected than randomly selected genes in the STRING PPI network. (A) Empirical distribution of the number of edges formed by 33 randomly selected genes. Green line represents the number of edges formed by the 33 CHD top genes from M-DATA. (B) Number of edges formed by CHD top genes, autism top genes from single-trait analyses and randomly selected genes, respectively.

Motivated by the observation from Fig 1, we develop a **N**etwork assisted model for **D**e novo **A**ssociation **T**est using protein-protein inter**A**ction information, named N-DATA, to leverage prior information of interactions among genes from the PPI network to boost statistical power in identifying risk genes for CHD. In the following, we first introduce the inference procedure for our model, and then demonstrate the performance of our method through simulation studies and real data applications.

## Methods

In this section, we introduce the statistical model for the proposed framework. The network information in the PPI database is represented by an undirected graph *G* = (*V, E*), where *V* = {1,…,*n*} is a set of *n* genes in the network, and *E* = {< *I, j* >: *i* and *j* are genes connected by the edges}. The degree of a gene *i* is defined as the number of direct neighbors (*N*_*i*_) for gene *i* in the network and denoted as *d*_*i*_ We denote the latent association status of gene *i* with a disease of interest, e.g., CHD, as *S*_*i*_, where *S*_*i*_ =1 if gene *i* is associated with the disease, *S*_*i*_ = −1 if gene *i* is not associated with the disease. *S =*{*S*_1_,..,*S*_*n*_} are the corresponding latent states for genes in *V* = {1, …, *n*}. The DNV count of the cohort is defined as *Y*_*i*_. To formalize the assumption that genes that have PPIs with risk genes are more likely to be risk genes, we apply a nearest neighbor Gibbs measure [33] to arrive at the following model:

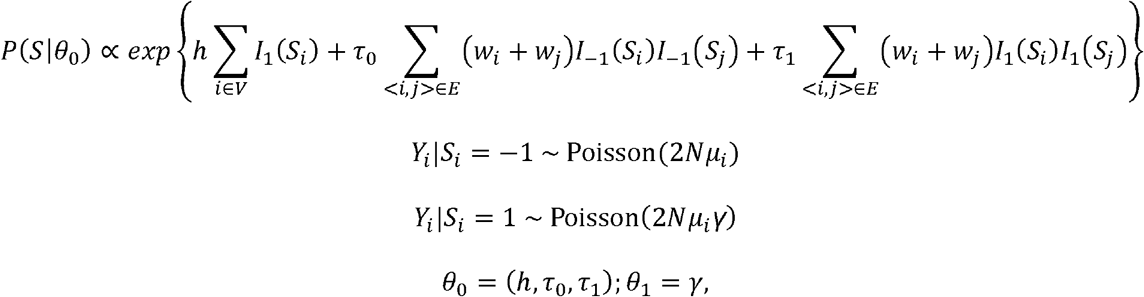

where *W*_*i*_ is the weight for gene *i* and will be chosen based on the characteristics of the network, *θ*_0_ = (*h, τ*_0_, *τ*_1_) are hyperparameters related to the network. Specifically, *h* determines the marginal distribution of *S*_*i*_ when all genes are independent i.e., 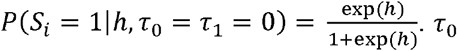. τ_0_ and τ_1_ characterize the prior weights of edges between non-associated genes and associated genes, respectively. *N* is the sample size of the case cohort, *μ*_*i*_ is the mutability of gene *i* estimated using the framework in Samocha et al. [34], and *θ*_1_ (*γ*) is the relative risk of the DNVs in the risk gene.

To reduce the computational burden from a fully Bayesian solution for maximizing the marginal likelihood, we propose an empirical Bayes method to estimate the parameters *θ*_0_ and *θ*_1_, and To reduce the computational burden from a fully Bayesian solution for maximizing the marginal the latent association status *S* by maximizing the pseudo conditional likelihood (PCLK) for *n* genes as follows

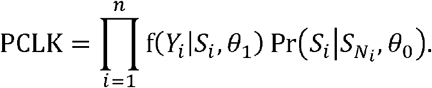

It has been shown that the estimator from the PCLK in a general Markov random field setting is consistent under mild regularity conditions [19] [35]. When maximizing the PCLK, we can estimate the hyperparameters *θ*_0,_ *θ*_1_ and latent status *S* iteratively.

We can obtain an empirical estimate for *θ*_0_ by maximizing 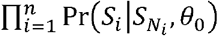, which is equivalent to maximizing the parameters in the following logistic regression model:

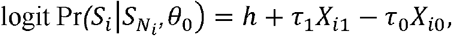

where 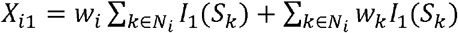 and 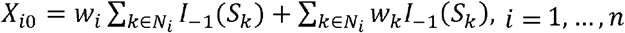 To make sure the estimated *θ*_0_ is finite, we can add a ridge penalty term 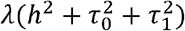 to the likelihood function to solve the maximization problem by the Newton-Raphson’s method [36].

We then update the latent status *S* by maximizing the PCLK using the iterative conditional mode method [35]. After we obtain the updated values *θ*_0_ and *S* we can estimate the hyperparameter *θ*_1_ by maximizing 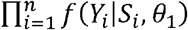 by using the following closed-form expression:

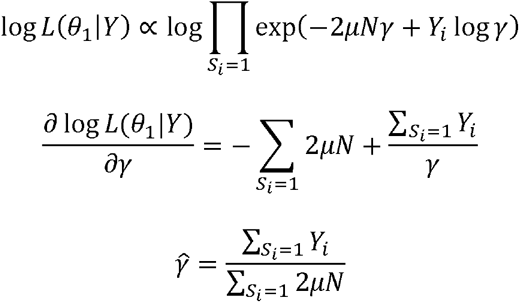

### Algorithm 1

Procedure for Parameter Estimation

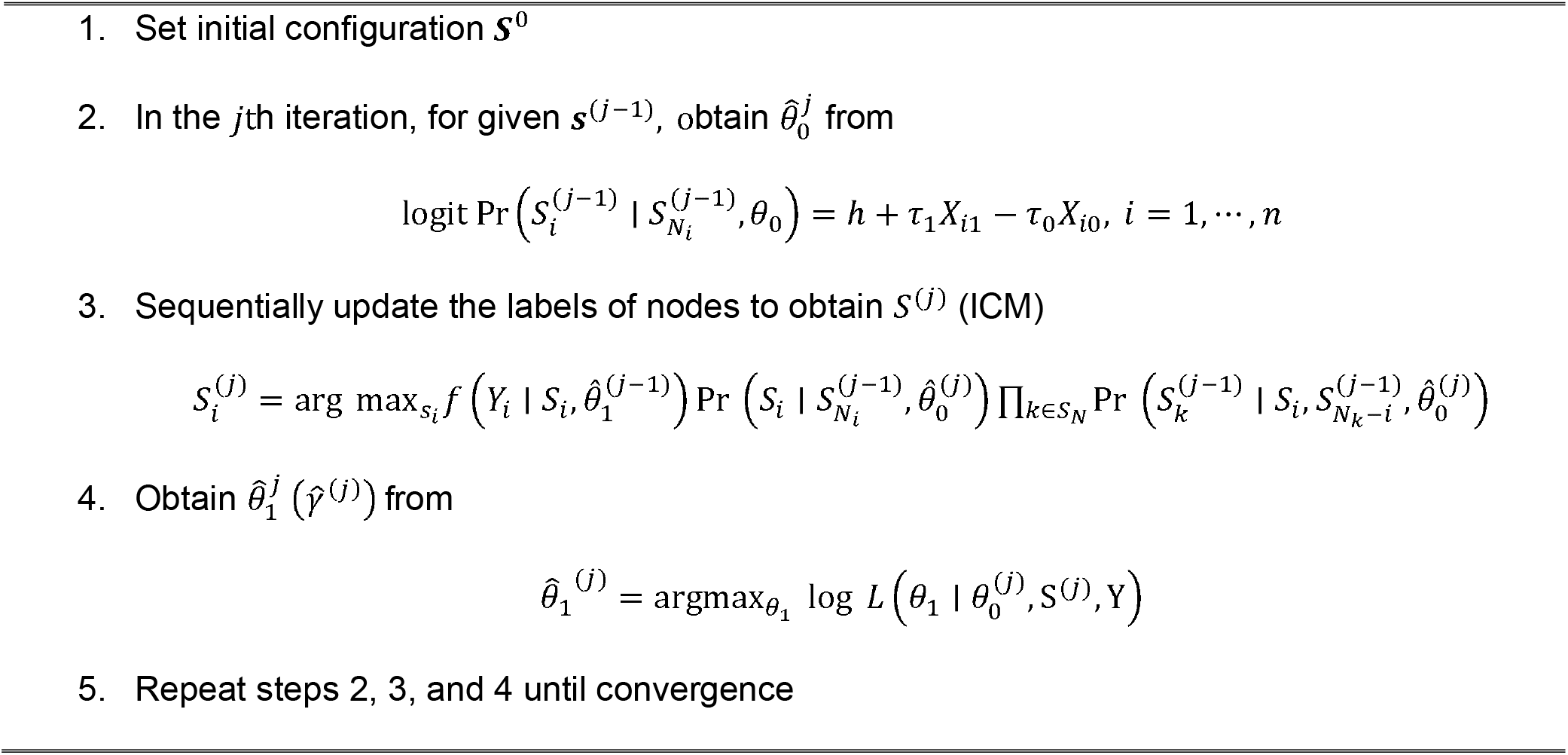

Finally, after we obtain the estimated 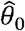 and 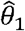, we use Gibbs sampling based on the conditional distribution 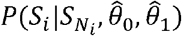 This method has been proved to be valid for multiple testing under dependence in a compound decision theoretic framework [37, 38]. Then, we can estimate the marginal posterior probability *q*_*i*_ = *P* (*S*_*i*_ = −1|*Y*).Let *q*_(*i*)_ be the sorted values of *q*_*i*_ in descending order. For each gene *i* the null hypothesis and alternative hypothesis are

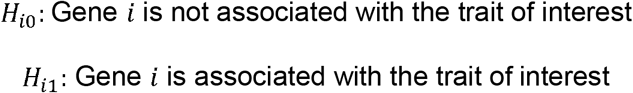

As shown by Jiang and Yu [39], the relationship between global FDR and local FDR (lfdr) is FDR = *E* (Ifdr|*Y* ∈ ℛ), where the rejection region ℛ is the set of *Y* such that the null hypothesis can be rejected based on a specific rejection criterion. To control the expected global FDR less than *α*, we propose the following procedure: let 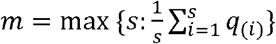, we reject all the null hypotheses corresponding to *H*_(1)_, …, *H*_(*m*)_.

## Results

### Simulation Studies

For DNV and network data, we considered similar settings as our real data. We randomly selected 2,000 genes, retrieved their mutability from the real data, and extracted the corresponding PPI network formed by these 2,000 genes. First, we simulated the true model with parameter values similar as those from real data analysis to see whether N-DATA can control FDR. Then, we evaluated the power under various settings of sample sizes *N* and relative risk parameter *γ*.

We set true network parameter *θ*_0_ as (−4, 0.2, 0) to make the total number of risk genes in the network of 2,000 random genes to be around 100. We varied the sample size *N* at 2,000, 5,000 and 10,000 to evaluate the performance of N-DATA in small, medium, and large WES cohorts, respectively. In addition, we varied *β* (log relative risk parameter *γ*) at 3.5, 4 and 4.5 to and 10,000 to investigate the performance of N-DATA around the burden estimated results from real data (In real data analysis, 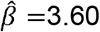) Each simulation setting was replicated 100 times. For Gibbs sampling-based inference, we used 2,000 MCMC iterations, and set the first 1,000 iterations as burn-ins. These numbers were chosen empirically based on the diagnostic plots for convergence. We report the performance under FDR threshold 0.05 in the main text (Fig 2) and FDR threshold 0.01 and 0.1 in Additional File 1.

**Fig 2.**
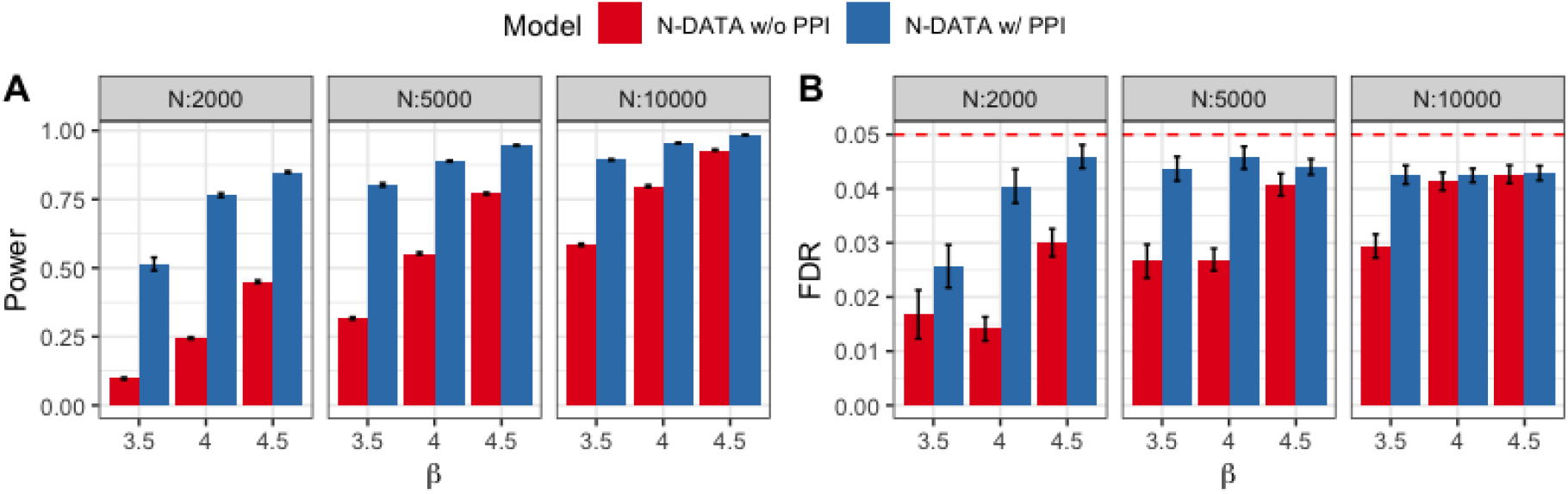
Comparison of N-DATA w/o and w/ PPI network models. (A) Power comparison of the two models. Three panels from left to right represent cohorts with small, medium, and large sample sizes, respectively. (B) FDR control for the two models. Red dash line represents the preset FDR threshold 0.05. Three panels from left to right represent cohorts with small, medium, and large sample sizes, respectively.

First, we compared the performance of N-DATA model with and without the PPI network as input. For N-DATA model without the PPI network, we assigned the weight of gene for inference. Both models controlled FDR well under all the settings. N-DATA model with PPI network had much better power than the model without PPI network when the sample size and were both low. When the sample size and were larger, the power of N-DATA without PPI network improved as expected.

Then, we compared the power of TADA-*De novo*, DAWN, and N-DATA using the same parameter settings. Hyperprior of TADA-*De novo* was estimated from the function *denovo*.*MOM* based on the recommendation from the authors [40]. Power of TADA was calculated based on TADA q-values. DAWN v1.0 was downloaded from http://www.compgen.pitt.edu/DAWN/DAWN_homepage.htm. We adapted the code of DAWN to use adjacency matrices of networks as its input. We used TADA-*De novo* p-values and PPI network as the input of DAWN. The parameter trim threshold was set to 4 based on the observation in our real data that, which corresponds to select the top 5 genes in TADA-*De novo* as fixed risk genes.

We compared the performance of TADA-*De novo*, TADA-*De novo* p-values + DAWN and N-DATA under different simulation settings. We reported the performance under FDR threshold 0.05 in the main text (Fig 3) and FDR threshold 0.01 and 0.1 in Additional File 1. We first checked if all three methods could control the global FDR when the threshold is 0.05. As discussed above, N-DATA controlled FDR well under all settings, while there was slight FDR inflation of TADA-*De novo* and DAWN under a couple of settings, especially when both and were very small or very large. N-DATA had the best power among the three methods under all settings. With an increase of and, the performance of TADA-*De novo* and DAWN also became better. For settings with FDR well controlled, DAWN performed better than TADA-*De novo*.

**Fig 3.**
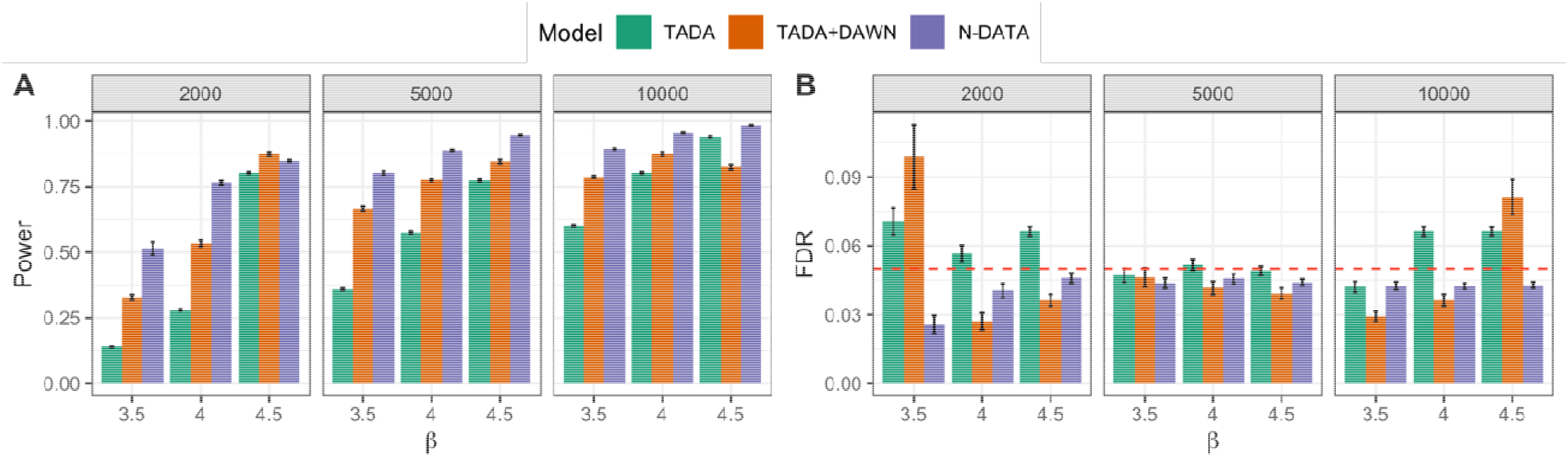
Comparison of TADA-*De novo*, TADA-*De novo* p-values + DAWN and N-DATA. Error bars represent standard errors estimated from 100 replications of simulation. (A) Power comparison of the three methods. Three panels from left to right represent cohorts with small, medium, and large sample sizes, respectively. (B) FDR control for the three methods. Red dash line represents the preset FDR threshold 0.05. Three panels from left to right represent cohorts with small, medium, and large sample sizes, respectively.

### Real Data Applications

We applied N-DATA to DNV data from 2,645 CHD trios reported in Jin et al [5]. We only considered damaging variants (loss of function (LoF) and deleterious missense (Dmis) variants by the MetaSVM algorithm) in our analysis as the number of non-deleterious variants is not expected to provide information to differentiate cases from controls biologically [41].

For network information, we first downloaded STRING v11.0 with medium edge likelihood via interface from STRINGdb package in R, and call this original network from STRING 𝒢_0_ obtained the curated list of known human CHD genes from Jin el al [5] and expanded the gene list by including additional candidate genes (FDR<0.1) from the single-trait analysis in our previous work [32]. Then, we extracted the subnetwork including the aforementioned gene list and the direct neighbors with likelihood score larger than 950 of those genes, and call this subnetwork 𝒢_1_. We only kept overlapping genes with our DNV data in 𝒢_1_ and called the final network used in our real application as 𝒢_2_. There were in total 1,818 genes and 21,534 edges in 𝒢_2_

To show that our method can leverage network information to boost risk gene identification, we applied our algorithm without using the network as an input. When there was no prior information from the network, we identified 18 significant genes with FDR<0.05. To include the network information from 𝒢_2_, we denote the degree of gene *i* in network 𝒢_2_ as *d*_*i*_, and let the weight in the prior as 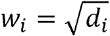 following Chen et al. [19]. After adding the network information from 𝒢_2_, we identified 46 genes with at least 1 DNV, and 26 genes harboring at least 2 DNVs with FDR<0.05 in the CHD cohort.

We also compared our results with TADA-*De novo* test [40] and DAWN [21, 42]. We downloaded the software of DAWN and adapted its code by substituting the adjacency matrix inferred from its Partial Neighborhood Selection algorithm to the adjacency matrix from network 𝒢_2_. The parameter settings were discussed in the simulation section. Specifically, we also compared two approaches to control the global FDR for TADA-*De novo* test (FDR q-values and p-values with FDR adjustment). FDR q-values given by TADA-De novo identified 25 significant genes, and p-values with FDR adjustment identified the same 25 genes. After integrating the p-values with the 𝒢_2_ network, 36 genes were identified by the adapted DAWN method. In total, N-DATA identified 325 genes with FDR<0.05. As some of the genes may be prioritized due to high prior probability, but did not have DNV count in the study cohort, we further filtered out genes without DNV and considered the 46 genes identified with FDR<0.05 and at least 1 DNV as the candidate genes. (Table 1)

**Table 1.**
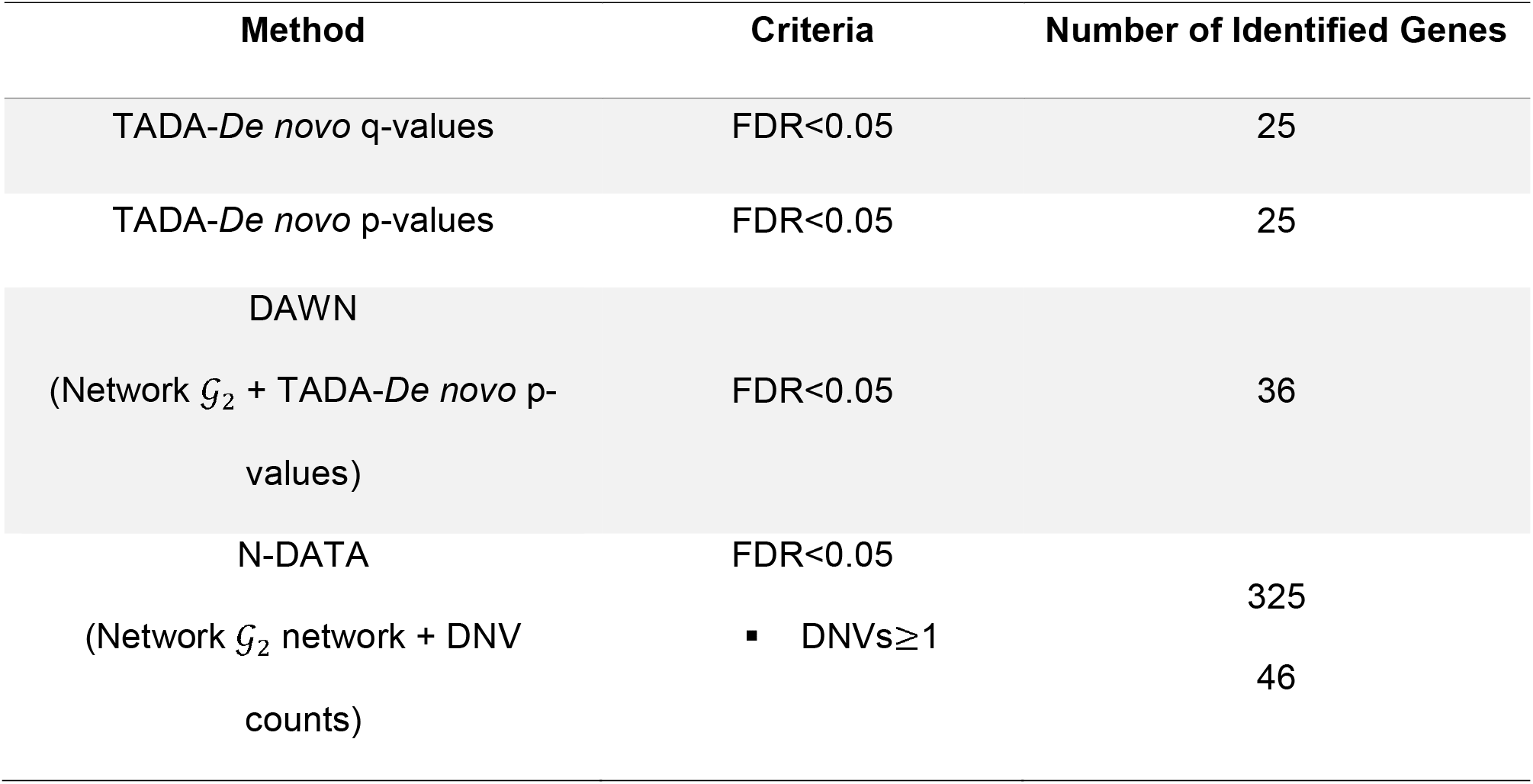
Comparison of N-DATA with other methods

We visualized the overlap of 253 known human CHD genes [5], genes that were identified by TADA-*De novo* q-values, DAWN, and N-DATA in Fig 4. Fig 4A shows the 325 genes identified by N-DATA, while Fig 4B shows the 46 genes with at least 1 DNV. From Fig 4B, N-DATA found all genes that can be identified by TADA, and identified 11 different genes compared with DAWN.

**Fig 4.**
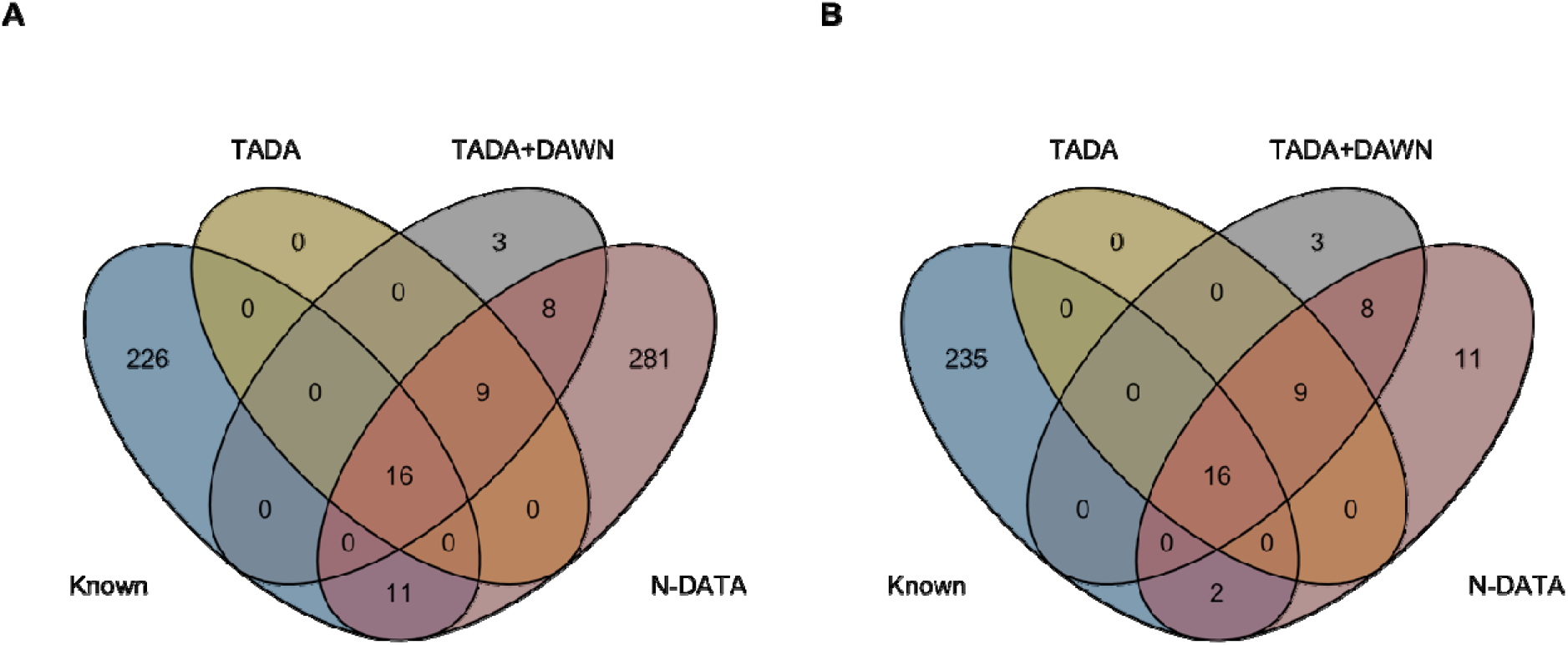
Venn diagram of known human CHD genes, TADA genes, DAWN genes and N-DATA genes. (A) Overlapping genes between known human CHD genes, TADA*-De novo*, TADA-*De novo* +DAWN and all 325 genes identified by N-DATA. (B) Overlapping genes between known human CHD genes, TADA*-De novo*, TADA-*De novo* +DAWN and 46 candidate genes identified by N-DATA.

Among the 46 genes, 18 are known human CHD genes and 35 are in the top 25% in mouse developing heart (HHE) at E14.5 [12]. Further, we calculated the overlap of the 46 N-DATA candidate genes, TADA genes, DAWN genes and 872 HHE genes that were analyzed in the 1,818 gene network. (Fig 5).

**Fig 5.**
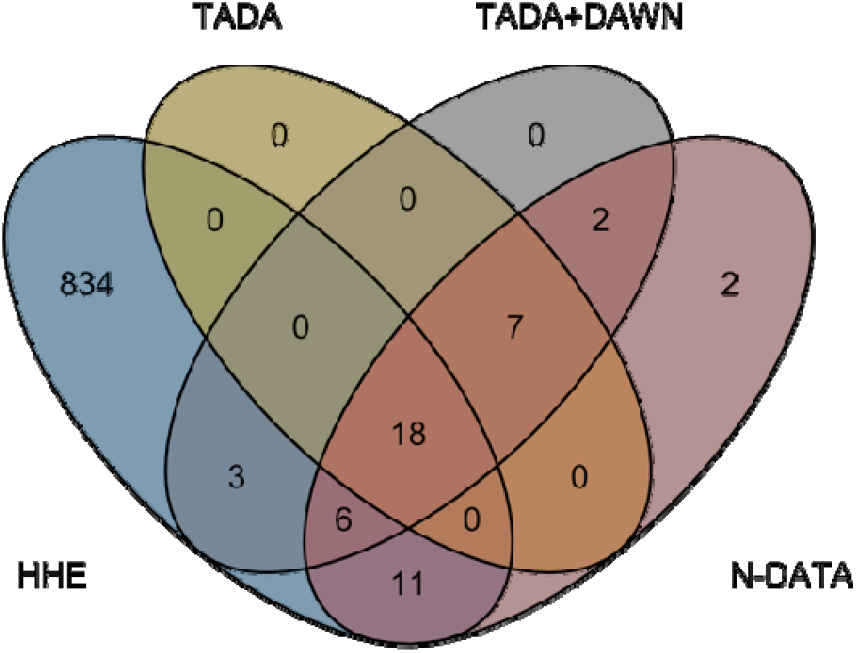
Venn diagram of HHE genes, TADA genes, DAWN genes and the 46 N-DATA candidate genes. N-DATA candidate genes identified 11 additional HHE genes compared with other methods.

We also visualized these 46 genes in the network to demonstrate that the PPI network information can help boost statistical power and provide biological interpretation.

Among the 46 candidate genes, *PTPN11, RAF1* and *RIT1* had 2 recurrent DNVs, and *CHD7, NOTCH1, NSD1* and *PYGL* also had recessive genotypes in the CHD cohort [5]. The 46 candidate genes form 4 clusters (Fig 6) in the 𝒢_2_ network. The biggest cluster includes seven known CHD genes *TBX5, KMT2D, PTPN11, SOS1, ACTB, NOTCH1*, and *PTEN*, which are involved in transcriptional regulation and the early cell growth or differentiation processes. The six new genes *SMAD2, KLF4, CTNNB1, CDC42, ITSN2*, and *WWTR1* also function in similar pathways and have varied implications in cardiac development. For instance, *KLF4* and *CTNNB1* have been implicated in cardiac cell differentiation [43]. *Cdc42* cardiomyocyte knock-out mice presented heart defects such as ventricular septum defects and thin ventricular walls [44]. *WWTR1* encodes a transcription regulator, which serves as an effector of Hippo pathway and regulates cardiac wall maturation in zebrafish [45].

**Fig 6.**
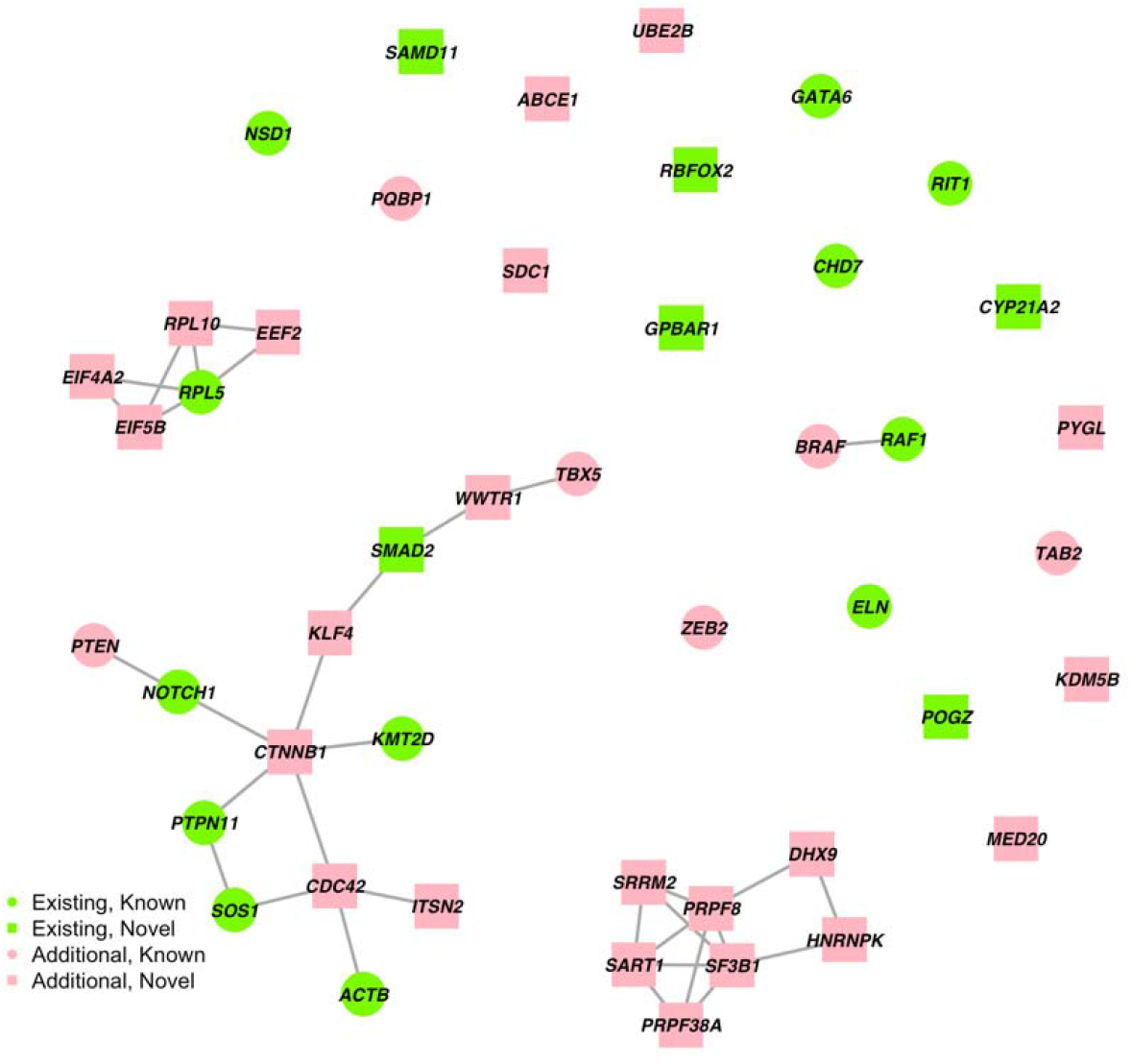
N-DATA model identified 46 candidate genes with at least 1 DNV. Green labels indicate the 18 genes identified when no network information was provided for N-DATA, and red labels indicate the additional 28 genes identified when the 𝒢 _2_ network was integrated. Circles indicate the 18 known human CHD genes, and squares indicate the 28 novel genes identified by N-DATA.

The second biggest cluster is constituted of 7 new genes, all of which are involved in mRNA splicing. Specifically, *SART1, SRRM2, PRPF38A, PRPF8*, and *SF3B1* are associated factors or components of spliceosome; *HNRNPK* encodes a pre-mRNA-binding protein; *DHX9* encodes an RNA helicase which promotes R-loop formation while RNA splicing is perturbed [46]. Alternative splicing plays an essential role in heart development, homeostasis, and disease pathogenesis. Mouse knockouts of multiple splice factors had impaired cardiogenesis [47]. *SF3B1*, specifically, has been shown to upregulate to induce heart disease in both human and mice [48]. Thus, though not fully investigated, DNVs in those mRNA splicing-related genes may contribute to CHD pathogenesis.

The third cluster contains genes involved in protein synthesis, includes the known gene *RPL5* and genes not previously associated with CHD (*EIF4, EIF*5, *EEF2*, and *RPL10). RPL5* and *RPL10* encode the ribosome subunits. Mutations in *RPL5* and other ribosomal genes can lead to multiple congenital anomalies, including CHD [49]. *EIF4* and *EIF5* encode translation initiation factors while *EEF2* encodes the elongation factor that regulate peptide chain elongation during protein synthesis. A recent study reported that the deficiency in ribosome associated NatA complex reduces ribosomal protein and subsequently impact cell development as a mechanism to cause CHD [50]. Thus, DNVs in the above genes may lead to CHD via impairment of protein synthesis.

The last cluster contains the known CHD genes *BRAF* and *RAF1*, both of which encode key kinases in Ras signaling and are related to Noonan syndrome with CHD as a common feature.

Among the un-clustered genes, six are identified after using the network information: *ABCE1, UBE2B, SDC1, PYGL, KDM5B, MED20. UBE2B* and *KDM5B*, encoding epigenetic modifiers, have shown suggestive evidence in cardiac development or CHD [51] [52] and might be potential CHD genes.

## Discussion

In this article, we have introduced a Bayesian framework to integrate PPI network information as the prior knowledge into DNV analysis for CHD. This approach adopts MRF to model the interactions among genes. We apply an empirical Bayes strategy to estimate parameters in the model and conduct statistical inference based on the posterior distribution sampled from a Gibbs sampler. The simulation studies and real data analysis on CHD suggest that the proposed method has improved power to identify risk genes over methods without integrating network information.

Our proposed framework is innovative for the following aspects. First, it can directly infer disease-associated genes using a network-based model without relying on summary statistics from other DNV association software. Second, it does not need to estimate hyperprior based on other sources compared to the existing pathway-based test for DNV data [17, 31]. Third, it does not require external expression data for the DNV cohort and uses the publicly available PPI database instead, which makes it more applicable to different diseases. This method will not only increase power in risk gene identification, but will also assist in biological interpretation by visualizing clusters of risk genes with functional relevance in the network.

However, there are some limitations in the current N-DATA model. In real application, it is important to conduct an initial analysis on the enrichment of top genes identified from *de novo* association test in the network like our motivating example. Another limitation is that the likelihood-based inference may suffer from local maxima [19]. Thus, we recommend to initiate the labels of genes from a known risk gene set or run with multiple starts. Also, we observe the Gibbs sampler tends to move around local maxima for some time before convergence. We suggest running at least 2,000 times of iterations and discard the first 1,000 iterations as burnins. In addition, we only considered damaging DNVs and assumed the relative risk parameter *³*, is the same across all genes in N-DATA, which may cause our model to lose power if it varies across variants with different functions (e.g., LoF and Dmis). Future studies may explore adding functional annotation of variants as a layer in the model to further improve statistical power.

## Conclusions

The topologic information in a pathway may be informative to identify functionally interrelated genes and help improve statistical power in DNV studies. Under the hypothesis that connected genes in PPI networks are more likely to share similar disease association status, we developed a novel statistical model that can leverage information from publicly available PPI databases. Through simulation studies under multiple settings, we proved our method can increase statistical power in identifying additional risk genes compared to methods without using the PPI network information. We then applied our method to the CHD DNV data, and then visualized the subnetwork of candidate genes to find potential functional gene clusters for CHD. Our results may shed new insight on the shared protein functionality among risk genes for CHD.

## Data Availability

All data produced in the present work are available online at https://github.com/JustinaXie/NDATA

## List of abbreviations

DNV: *de novo* variant
CHD: congenital heart disease
WES: Whole Exome Sequencing
PPI: protein-protein Interaction
FDR: false discovery rate
MRF: Markov Random Field
GWAS: Genome-Wide Association Studies

## Acknowledgements

We thank Jin et al. [5] for sharing the *de novo* variant data of CHD. We thank Andrew Xu and Dr. Min Chen for discussions on coding, and Ziyu Jiang for discussions on diagnostic for Bayesian inference.

## Funding

This work was supported in part by NIH grant R03HD100883-01A1 (Y.X. and H.Z.) and R01GM134005-01A1 (W.J., H.L., and H.Z.). The funders had no role in study design, data collection and analysis, decision to publish, or preparation of the manuscript.

## Ethics Declaration

### Ethics approval and consent to participate

This manuscript is a mainly methodology paper. The data used in this manuscript is all public available. The research has no procedure related to collecting human subject data.

## Consent for publication

Not applicable.

## Competing interests

The authors declare that they have no competing interests.

## Supplementary Information

Additional File 1. Supplementary figures 1-4

## Notes

### Competing Interest Statement

The authors have declared no competing interest.

### Funding Statement

This study was funded in part by NIH grant R03HD100883-01A1 (Y.X. and H.Z.) and R01GM134005-01A1 (W.J., H.L., and H.Z.).

### Author Declarations

This manuscript is a mainly methodology paper. The data used in this manuscript are all public available. The research has no procedure related to collecting human subject data.2,645 CHD data can be downloaded from the supplement of PMID28991257 (https://static-content.springer.com/esm/art%3A10.1038%2Fng.3970/MediaObjects/41588_2017_BFng3970_MOESM3_ESM.xlsx).

## References

1. Zaidi S, Choi M, Wakimoto H, Ma L, Jiang J, Overton JD, Romano-Adesman A, Bjornson RD, Breitbart RE, Brown KK, et al: De novo mutations in histone-modifying genes in congenital heart disease. Nature 2013, 498:220–223.

2. Postma AV, Bezzina CR, Christoffels VM: Genetics of congenital heart disease: the contribution of the noncoding regulatory genome. Journal of Human Genetics 2016, 61:13–19.

3. Sevim Bayrak C, Zhang P, Tristani-Firouzi M, Gelb BD, Itan Y: De novo variants in exomes of congenital heart disease patients identify risk genes and pathways. Genome Med 2020, 12:9.

4. Diab NS, Barish S, Dong W, Zhao S, Allington G, Yu X, Kahle KT, Brueckner M, Jin SC: Molecular Genetics and Complex Inheritance of Congenital Heart Disease. Genes (Basel) 2021, 12.

5. Jin SC, Homsy J, Zaidi S, Lu Q, Morton S, DePalma SR, Zeng X, Qi H, Chang W, Sierant MC, et al: Contribution of rare inherited and de novo variants in 2,871 congenital heart disease probands. Nat Genet 2017, 49:1593–1601.

6. Zaidi S, Brueckner M: Genetics and Genomics of Congenital Heart Disease. Circ Res 2017, 120:923–940.

7. Glessner JT, Bick AG, Ito K, Homsy J, Rodriguez-Murillo L, Fromer M, Mazaika E, Vardarajan B, Italia M, Leipzig J, et al: Increased frequency of de novo copy number variants in congenital heart disease by integrative analysis of single nucleotide polymorphism array and exome sequence data. Circ Res 2014, 115:884–896.

8. Soemedi R, Wilson IJ, Bentham J, Darlay R, Töpf A, Zelenika D, Cosgrove C, Setchfield K, Thornborough C, Granados-Riveron J, et al: Contribution of global rare copy-number variants to the risk of sporadic congenital heart disease. Am J Hum Genet 2012, 91:489–501.

9. Pierpont ME, Brueckner M, Chung WK, Garg V, Lacro RV, McGuire AL, Mital S, Priest JR, Pu WT, Roberts A, et al: Genetic Basis for Congenital Heart Disease: Revisited: A Scientific Statement From the American Heart Association. Circulation 2018, 138:e653–e711.

10. Teer JK, Mullikin JC: Exome sequencing: the sweet spot before whole genomes. Human Molecular Genetics 2010, 19:R145–R151.

11. Rabbani B, Tekin M, Mahdieh N: The promise of whole-exome sequencing in medical genetics. Journal of Human Genetics 2014, 59:5–15.

12. Homsy J, Zaidi S, Shen Y, Ware JS, Samocha KE, Karczewski KJ, DePalma SR, McKean D, Wakimoto H, Gorham J, et al: De novo mutations in congenital heart disease with neurodevelopmental and other congenital anomalies. Science 2015, 350:1262–1266.

13. Richter F, Morton SU, Kim SW, Kitaygorodsky A, Wasson LK, Chen KM, Zhou J, Qi H, Patel N, DePalma SR: Genomic analyses implicate noncoding de novo variants in congenital heart disease. Nature genetics 2020, 52:769–777.

14. Watkins WS, Hernandez EJ, Wesolowski S, Bisgrove BW, Sunderland RT, Lin E, Lemmon G, Demarest BL, Miller TA, Bernstein D: De novo and recessive forms of congenital heart disease have distinct genetic and phenotypic landscapes. Nature communications 2019, 10:1–12.

15. Sifrim A, Hitz M-P, Wilsdon A, Breckpot J, Turki SHA, Thienpont B, McRae J, Fitzgerald TW, Singh T, Swaminathan GJ, et al: Distinct genetic architectures for syndromic and nonsyndromic congenital heart defects identified by exome sequencing. Nature Genetics 2016, 48:1060–1065.

16. Sifrim A, Hitz MP, Wilsdon A, Breckpot J, Turki SH, Thienpont B, McRae J, Fitzgerald TW, Singh T, Swaminathan GJ, et al: Distinct genetic architectures for syndromic and nonsyndromic congenital heart defects identified by exome sequencing. Nat Genet 2016, 48:1060–1065.

17. Nguyen TH, He X, Brown RC, Webb BT, Kendler KS, Vladimirov VI, Riley BP, Bacanu SA: DECO: a framework for jointly analyzing de novo and rare case/control variants, and biological pathways. Brief Bioinform 2021.

18. Lee I, Blom UM, Wang PI, Shim JE, Marcotte EM: Prioritizing candidate disease genes by network-based boosting of genome-wide association data. Genome Res 2011, 21:1109–1121.

19. Chen M, Cho J, Zhao H: Incorporating biological pathways via a Markov random field model in genome-wide association studies. PLoS Genet 2011, 7:e1001353.

20. Hou L, Chen M, Zhang CK, Cho J, Zhao H: Guilt by rewiring: gene prioritization through network rewiring in genome wide association studies. Hum Mol Genet 2014, 23:2780–2790.

21. Liu L, Lei J, Roeder K: Network assisted analysis to reveal the genetic basis of autism. The Annals of Applied Statistics 2015, 9:1571-1600, 1530.

22. Nguyen HT, Bryois J, Kim A, Dobbyn A, Huckins LM, Munoz-Manchado AB, Ruderfer DM, Genovese G, Fromer M, Xu X, et al: Integrated Bayesian analysis of rare exonic variants to identify risk genes for schizophrenia and neurodevelopmental disorders. Genome Med 2017, 9:114.

23. Nguyen T-H, Dobbyn A, Brown RC, Riley BP, Buxbaum JD, Pinto D, Purcell SM, Sullivan PF, He X, Stahl EA: mTADA is a framework for identifying risk genes from de novo mutations in multiple traits. Nature Communications 2020, 11:2929.

24. Oughtred R, Stark C, Breitkreutz BJ, Rust J, Boucher L, Chang C, Kolas N, O’Donnell L, Leung G, McAdam R, et al: The BioGRID interaction database: 2019 update. Nucleic Acids Res 2019, 47:D529–d541.

25. Orchard S, Ammari M, Aranda B, Breuza L, Briganti L, Broackes-Carter F, Campbell NH, Chavali G, Chen C, del-Toro N, et al: The MIntAct project--IntAct as a common curation platform for 11 molecular interaction databases. Nucleic Acids Res 2014, 42:D358–363.

26. Salwinski L, Miller CS, Smith AJ, Pettit FK, Bowie JU, Eisenberg D: The Database of Interacting Proteins: 2004 update. Nucleic Acids Res 2004, 32:D449–451.

27. Licata L, Briganti L, Peluso D, Perfetto L, Iannuccelli M, Galeota E, Sacco F, Palma A, Nardozza AP, Santonico E, et al: MINT, the molecular interaction database: 2012 update. Nucleic Acids Res 2012, 40:D857–861.

28. Mishra GR, Suresh M, Kumaran K, Kannabiran N, Suresh S, Bala P, Shivakumar K, Anuradha N, Reddy R, Raghavan TM, et al: Human protein reference database--2006 update. Nucleic Acids Res 2006, 34:D411–414.

29. Szklarczyk D, Gable AL, Lyon D, Junge A, Wyder S, Huerta-Cepas J, Simonovic M, Doncheva NT, Morris JH, Bork P, et al: STRING v11: protein-protein association networks with increased coverage, supporting functional discovery in genome-wide experimental datasets. Nucleic Acids Res 2019, 47:D607–d613.

30. Nguyen TH, Dobbyn A, Brown RC, Riley BP, Buxbaum JD, Pinto D, Purcell SM, Sullivan PF, He X, Stahl EA: mTADA is a framework for identifying risk genes from de novo mutations in multiple traits. Nat Commun 2020, 11:2929.

31. Nguyen HT, Dobbyn A, Charney AW, Bryois J, Kim A, Mcfadden W, Skene NG, Huckins LM, Wang W, Ruderfer DM, et al: Integrative analysis of rare variants and pathway information shows convergent results between immune pathways, drug targets and epilepsy genes. bioRxiv 2018:410100.

32. Xie Y, Li M, Dong W, Jiang W, Zhao H: M-DATA: A statistical approach to jointly analyzing de novo mutations for multiple traits. PLoS Genet 2021, 17:e1009849.

33. Kindermann R: Markov random fields and their applications. American mathematical society 1980.

34. Samocha KE, Robinson EB, Sanders SJ, Stevens C, Sabo A, McGrath LM, Kosmicki JA, Rehnström K, Mallick S, Kirby A, et al: A framework for the interpretation of de novo mutation in human disease. Nat Genet 2014, 46:944–950.

35. Besag J: On the statistical analysis of dirty pictures. Journal of the Royal Statistical Society: Series B (Methodological) 1986, 48:259–279.

36. Le Cessie S, Van Houwelingen JC: Ridge estimators in logistic regression. Journal of the Royal Statistical Society: Series C (Applied Statistics) 1992, 41:191–201.

37. Sun W, Tony Cai T: Large-scale multiple testing under dependence. Journal of the Royal Statistical Society: Series B (Statistical Methodology) 2009, 71:393–424.

38. Li H, Wei Z, Maris J: A hidden Markov random field model for genome-wide association studies. Biostatistics 2010, 11:139–150.

39. Jiang W, Yu W: Controlling the joint local false discovery rate is more powerful than meta-analysis methods in joint analysis of summary statistics from multiple genome-wide association studies. Bioinformatics 2016, 33:500–507.

40. He X, Sanders SJ, Liu L, De Rubeis S, Lim ET, Sutcliffe JS, Schellenberg GD, Gibbs RA, Daly MJ, Buxbaum JD, et al: Integrated model of de novo and inherited genetic variants yields greater power to identify risk genes. PLoS Genet 2013, 9:e1003671.

41. Li M: Gene-based Association Analysis for Genome-wide Association and Whole-exome Sequencing Studies. Yale University, Biostatistics; 2020.

42. Liu L, Lei J, Sanders SJ, Willsey AJ, Kou Y, Cicek AE, Klei L, Lu C, He X, Li M, et al: DAWN: a framework to identify autism genes and subnetworks using gene expression and genetics. Mol Autism 2014, 5:22.

43. Kami D, Kitani T, Kawasaki T, Gojo S: Cardiac mesenchymal progenitors differentiate into adipocytes via Klf4 and c-Myc. Cell death & disease 2016, 7:e2190–e2190.

44. Liu Y, Wang J, Li J, Wang R, Tharakan B, Zhang SL, Tong CW, Peng X: Deletion of Cdc42 in embryonic cardiomyocytes results in right ventricle hypoplasia. Clinical and translational medicine 2017, 6:40–40.

45. Lai JKH, Collins MM, Uribe V, Jiménez-Amilburu V, Günther S, Maischein HM, Stainier DYR: The Hippo pathway effector Wwtr1 regulates cardiac wall maturation in zebrafish. Development 2018, 145.

46. Chakraborty P, Huang JTJ, Hiom K: DHX9 helicase promotes R-loop formation in cells with impaired RNA splicing. Nat Commun 2018, 9:4346.

47. Zahr HC, Jaalouk DE: Exploring the Crosstalk Between LMNA and Splicing Machinery Gene Mutations in Dilated Cardiomyopathy. Front Genet 2018, 9:231.

48. van den Hoogenhof MM, Pinto YM, Creemers EE: RNA Splicing: Regulation and Dysregulation in the Heart. Circ Res 2016, 118:454–468.

49. Gazda HT, Sheen MR, Vlachos A, Choesmel V, O’Donohue MF, Schneider H, Darras N, Hasman C, Sieff CA, Newburger PE, et al: Ribosomal protein L5 and L11 mutations are associated with cleft palate and abnormal thumbs in Diamond-Blackfan anemia patients. Am J Hum Genet 2008, 83:769–780.

50. Ward T, Tai W, Morton S, Impens F, Van Damme P, Van Haver D, Timmerman E, Venturini G, Zhang K, Jang MY, et al: Mechanisms of Congenital Heart Disease Caused by NAA15 Haploinsufficiency. Circ Res 2021, 128:1156–1169.

51. Robson A, Makova SZ, Barish S, Zaidi S, Mehta S, Drozd J, Jin SC, Gelb BD, Seidman CE, Chung WK, et al: Histone H2B monoubiquitination regulates heart development via epigenetic control of cilia motility. Proc Natl Acad Sci U S A 2019, 116:14049–14054.

52. Audain E, Wilsdon A, Breckpot J, Izarzugaza JMG, Fitzgerald TW, Kahlert AK, Sifrim A, Wünnemann F, Perez-Riverol Y, Abdul-Khaliq H, et al: Integrative analysis of genomic variants reveals new associations of candidate haploinsufficient genes with congenital heart disease. PLoS Genet 2021, 17:e1009679.

