## Additional File 1 for "Network Assisted Analysis of *De Novo* Variants Using Protein-Protein Interaction Information Identified 46 Candidate Genes for Congenital Heart Disease"

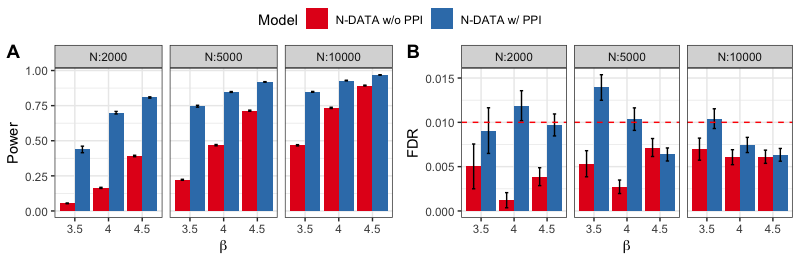

**Fig S1**. **Comparison of N-DATA w/o and w/ PPI network models**. (A) Power comparison of the two models. Three panels from left to right represent cohorts with small, medium, and large sample sizes, respectively. (B) FDR control for the two models. Red dash line represents the preset FDR threshold 0.01. Three panels from left to right represent cohorts with small, medium, and large sample sizes, respectively.

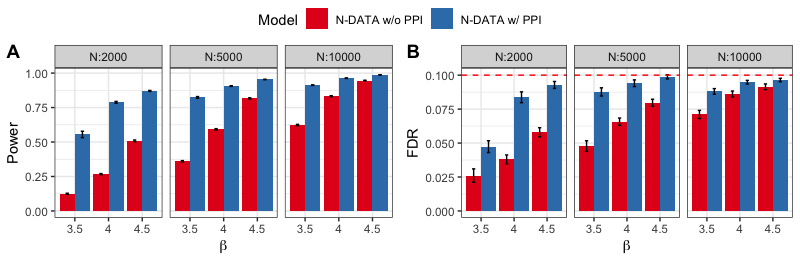

**Fig S2**. **Comparison of N-DATA w/o and w/ PPI network models**. (A) Power comparison of the two models. Three panels from left to right represent cohorts with small, medium, and large sample sizes, respectively. (B) FDR control for the two models. Red dash line represents the preset FDR threshold 0.1. Three panels from left to right represent cohorts with small, medium, and large sample sizes, respectively.

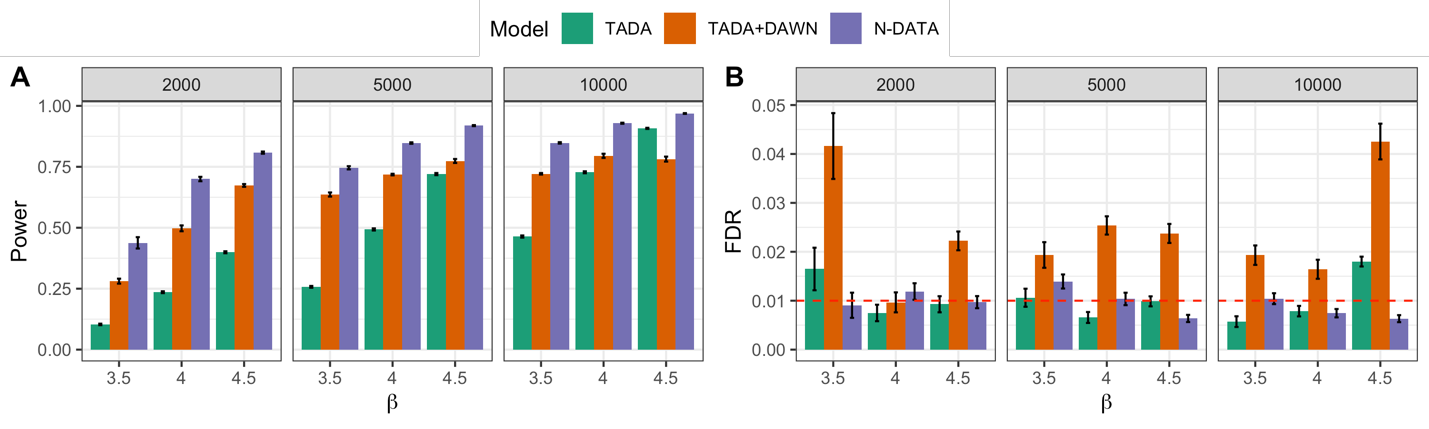

**Fig S3**. **Comparison of TADA-*De novo*, TADA-*De novo* p-values + DAWN and N-DATA**. Error bars represent standard errors estimated from 100 replications of simulation. (A) Power comparison of the three methods. Three panels from left to right represent cohorts with small, medium, and large sample sizes, respectively. (B) FDR control for the three methods. Red dash line represents the preset FDR threshold 0.01. Three panels from left to right represent cohorts with small, medium, and large sample sizes, respectively.

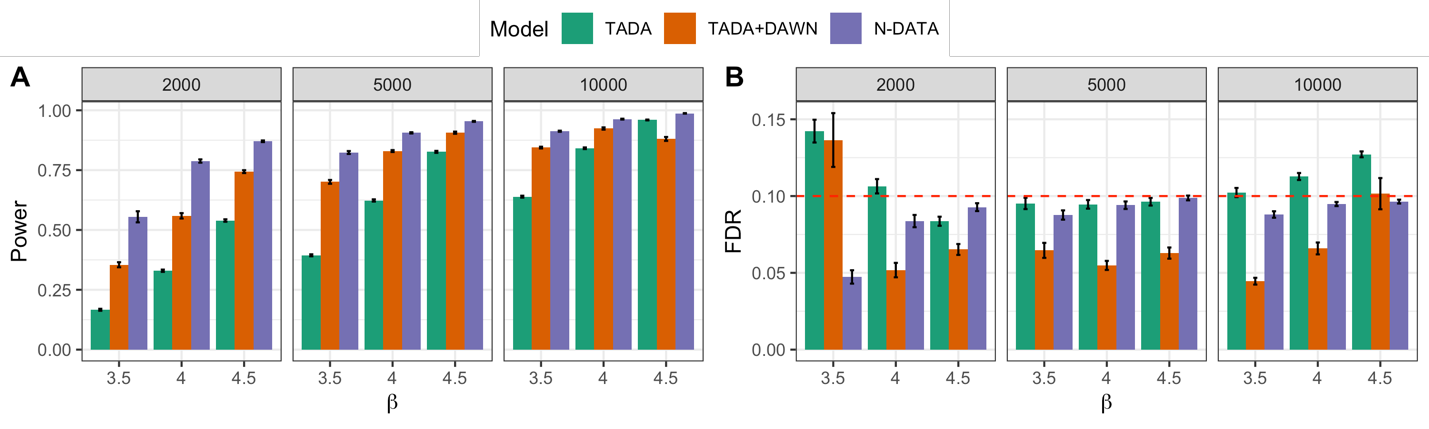

**Fig S4**. **Comparison of TADA-*De novo*, TADA-*De novo* p-values + DAWN and N-DATA**. Error bars represent standard errors estimated from 100 replications of simulation. (A) Power comparison of the three methods. Three panels from left to right represent cohorts with small, medium, and large sample sizes, respectively. (B) FDR control for the three methods. Red dash line represents the preset FDR threshold 0.1. Three panels from left to right represent cohorts with small, medium, and large sample sizes, respectively.
